# Socio-demographic and behavioral factors influencing the utilization of community post interventions for HIV testing services by men (≥18 years) in Kericho County, Kenya

**DOI:** 10.64898/2026.09.03.26362133

**Authors:** Lilian N. Kong’ani, Jackline Mosinya Nyaberi, Kenneth Ngure

## Abstract

Men continue to test for HIV less often than women in many settings, including Kenya. Community-based HIV testing may reduce access barriers, but evidence on factors associated with men’s use of community-post services remains limited. This analytical cross-sectional mixed-methods study examined socio-demographic and behavioral factors associated with use of community posts for HIV testing among adult men in Kericho County, Kenya. Quantitative data were collected using self-administered questionnaires at two community posts and two facility-based Comprehensive Care Centres. The Kobo export contained 438 questionnaire records; 41 pretest records were excluded in line with the study protocol, leaving 397 records for analysis. Community-post use in the preceding year was reported by 208 of 396 respondents with complete outcome data (52.5%). In adjusted analysis, community-post use was higher among employed men (AOR 2.94, 95% CI 1.48–5.82; p=0.002) and men engaged in business (AOR 2.56, 95% CI 1.15–5.69; p=0.021) than among unemployed/other men. Age, marital status, religion, education and monthly income were not independently associated with community-post use. In the behavioral model, respondents who were unwilling or unsure about participating in future community HIV-testing campaigns had lower odds of community-post use (AOR 0.20, 95% CI 0.10–0.39; p<0.001). Frequent travel or migration (AOR 0.19, 95% CI 0.06–0.65; p=0.008) and unprotected sex (AOR 0.54, 95% CI 0.34–0.87; p=0.012) were also associated with lower odds of community-post use after adjustment. These findings identify occupational and selected behavioral factors associated with community-post use among the men studied; the cross-sectional design does not establish causality.

## Introduction

HIV remains a major global public health challenge. In 2023, an estimated 39.9 million people were living with HIV, 1.3 million people acquired HIV, and 630,000 people died from AIDS-related illnesses [1]. Men continue to have poorer engagement across parts of the HIV testing and treatment cascade. In Kenya, the 2022 Demographic and Health Survey reported that 73% of men had ever been tested for HIV and received their results, compared with 85% of women [2].

Community-based HIV testing services bring testing closer to populations that may be less likely to attend health facilities. Evidence from systematic reviews and implementation studies shows that community-based and peer-led approaches can increase testing access, including among men, and can reach first-time testers and people who are unaware of their HIV status [3–6]. Kenyan evidence similarly identifies facility location, waiting time, clinic hours, confidentiality, stigma and perceived provider attitudes as factors that shape men’s testing decisions [7].

Socio-demographic and behavioral characteristics can further influence HIV testing. Studies in eastern and sub-Saharan Africa have identified variation in testing by age, education, residence, wealth, marital status and other individual or contextual factors [8,9]. Sexual risk, peer or community engagement, and perceived accessibility of testing may also influence whether and where men test [5,7,10]. These relationships are context dependent and require local evidence to guide differentiated testing strategies.

In Kericho County, community posts were introduced as decentralized points for HIV testing and related services. However, evidence on the factors associated with men’s utilization of these community-post services has been limited. This study therefore aimed to determine the proportion of adult men utilizing HIV testing services and to establish the socio-demographic and behavioral factors associated with utilization of community-post HIV testing interventions in Kericho County, Kenya.

## Materials and Methods

### Study design and setting

An analytical cross-sectional mixed-methods study was conducted in Kericho County, Kenya. Participants were recruited from two community posts (Nyagacho and Brooke) and two health facilities with Comprehensive Care Centres (CCCs): Kericho County Referral Hospital and Londiani Sub-County Hospital. The study covered the period from 1 July 2024 to 31 July 2025. The quantitative component compared patterns of HIV testing utilization across the two service-delivery settings, while qualitative interviews explored contextual factors influencing men’s testing decisions.

### Study population and sampling

The quantitative study population comprised men aged 18 years and above recruited at Nyagacho and Brooke community posts and at Kericho County Referral Hospital and Londiani Sub-County Hospital. Eligible participants were adult men who met the study eligibility criteria and provided informed consent. The questionnaire was pretested before the main study. The Kobo export supplied for verification contained 438 questionnaire records. Consistent with the study protocol and the analytic denominator used in the thesis, 41 pretest records were excluded, leaving 397 records in the main analytic dataset. One participant had a missing response to the community-post utilization outcome and was excluded only from analyses requiring that outcome.

### Data collection

Quantitative data were collected using self-administered questionnaires and HIV testing service record abstraction. The questionnaire captured socio-demographic characteristics, knowledge and utilization of community posts, testing frequency, perceived importance of HIV testing, discussion of testing with social contacts, selected HIV-risk behaviors, mobility, alcohol or drug use, and peer influence. Instruments were pretested in Kisumu County using approximately 10% of the planned sample. Qualitative data were collected through key informant interviews and focus group discussions using semi-structured guides. The interviews explored perceived use of community posts relative to facility-based testing, accessibility and convenience, confidentiality and stigma, socio-demographic and behavioral influences, provider attitudes and service delivery, and suggested improvements to community-based HIV testing services.

### Outcome and explanatory variables

The primary quantitative outcome was self-reported use of a community post for HIV testing during the preceding year, derived from the multiple-response item asking where the participant had accessed HIV testing services. The community-post option was coded as yes or no. Socio-demographic explanatory variables included age, marital status, religion, occupation, education and monthly household income. Behavioral variables included perceived importance of HIV testing, discussion of HIV testing with friends, family or peers, willingness to participate in future community HIV-testing campaigns, multiple sexual partnerships, unprotected sex, injection-drug use, frequent travel or migration, alcohol or drug use, and peer influence. For multivariable analysis, occupation was grouped as employed, farmer, business, and unemployed/other; education was grouped as primary, secondary and tertiary; and the two oldest age categories were combined as 56 years and above because of sparse outcome counts.

### Statistical and qualitative analysis

Quantitative data were analysed in STATA version 14. Frequencies and percentages were used to summarize categorical variables. Multivariable logistic regression was used to estimate adjusted odds ratios (AORs), 95% confidence intervals (CIs) and p-values for factors associated with community-post utilization. The socio-demographic model included age, marital status, religion, occupation, education and monthly income. The behavioral model included frequency of discussion of HIV testing, perceived importance of testing, willingness to participate in future campaigns, multiple sexual partnerships, unprotected sex, injection-drug use, frequent travel or migration, alcohol or drug use, and peer influence. The mutually exclusive ‘no risk behavior’ option was not entered simultaneously with the individual risk-behavior indicators to avoid redundant coding. Statistical significance was assessed at p<0.05. Qualitative data were analysed thematically using NVivo 14. Themes were organized around accessibility and convenience, confidentiality and stigma, social and behavioral influences, and provider/service-delivery factors, and were used to interpret the quantitative findings without converting qualitative perceptions into quantitative estimates.

### Ethical considerations

Ethical approval was obtained from the Masinde Muliro University of Science and Technology Institutional Research and Ethics Committee (MMUST IREC), Ref: MMU/COR:40312 Vol 6(01), on 2 April 2025. A data-collection authorization letter was subsequently obtained from Jomo Kenyatta University of Agriculture and Technology. A national research permit, Ref. 277884, was obtained from the National Commission for Science, Technology and Innovation (NACOSTI) on 26 June 2025. Participation was voluntary, and written informed consent was obtained from participants before data collection. Confidentiality was maintained throughout data collection and analysis.

## Results

### Participant characteristics

The main analytic dataset included 397 men. The largest age group was 26–35 years (146/397; 36.8%), followed by 18–25 years (127/397; 32.0%). Most participants were unmarried (229/397; 57.7%) and Christian (375/397; 94.5%). The most common occupations were casual labour (114/397; 28.7%), farming (76/397; 19.1%), business (71/397; 17.9%) and formal employment (67/397; 16.9%). Secondary education was reported by 195/397 (49.1%), while 150/397 (37.8%) had TVET, college or university education. More than half (214/397; 53.9%) reported monthly household income of KES 10,000 or less.

### HIV testing utilization

All 397 participants reported at least one HIV test in the preceding year. Community-post testing during that period was reported by 208 of 396 participants with complete responses to the service-location item (52.5%); one response was missing. Government facilities were reported by 165/397 (41.6%), private hospitals or pharmacies by 48/397 (12.1%), mobile clinics or outreach services by 38/397 (9.6%), and faith-based organizations by 28/397 (7.1%). Because participants could select more than one testing location, these percentages are not mutually exclusive. Awareness of community posts providing HIV testing, combining the equivalent item across the two Kobo form versions, was reported by 318/397 (80.1%). Most men reported testing twice in the preceding year (237/397; 59.7%), followed by once (107/397; 27.0%), three times (45/397; 11.3%) and more than three times (8/397; 2.0%).

**Table 1.** Awareness, HIV-testing frequency and reported testing locations in the verified analytic dataset.

| **Indicator** | **Category** | **n/N (%)** |
| --- | --- | --- |
| Awareness of community posts | Yes | 318/397 (80.1) |
| Awareness of community posts | No | 79/397 (19.9) |
| Testing frequency | Once | 107/397 (27.0) |
| Testing frequency | Twice | 237/397 (59.7) |
| Testing frequency | Thrice | 45/397 (11.3) |
| Testing frequency | >3 times | 8/397 (2.0) |
| Testing location* | Community post | 208/396 (52.5) |
| Testing location* | Government facility | 165/397 (41.6) |
| Testing location* | Private hospital/pharmacy | 48/397 (12.1) |
| Testing location* | Mobile/outreach | 38/397 (9.6) |
| Testing location* | Faith-based organization | 28/397 (7.1) |

Participants could report more than one place of HIV testing in the preceding year. The raw-data verification therefore treats each testing location as a separate multiple-response indicator rather than as mutually exclusive categories.

### Socio-demographic factors associated with community-post utilization

In the adjusted socio-demographic model (n=396), occupation was the only socio-demographic domain independently associated with community-post utilization. Compared with men classified as unemployed/other, employed men had nearly three times the odds of reporting community-post use (AOR 2.94, 95% CI 1.48–5.82; p=0.002), while men engaged in business had higher odds of use (AOR 2.56, 95% CI 1.15–5.69; p=0.021). Farming was not independently associated with utilization (AOR 1.32, 95% CI 0.58–3.01; p=0.511). Age, marital status, religion, education and monthly household income were not independently associated with community-post use in this model.

In the adjusted model, age, marital status, religion, education and monthly household income were not independently associated with community-post utilization.

**Table 2.**
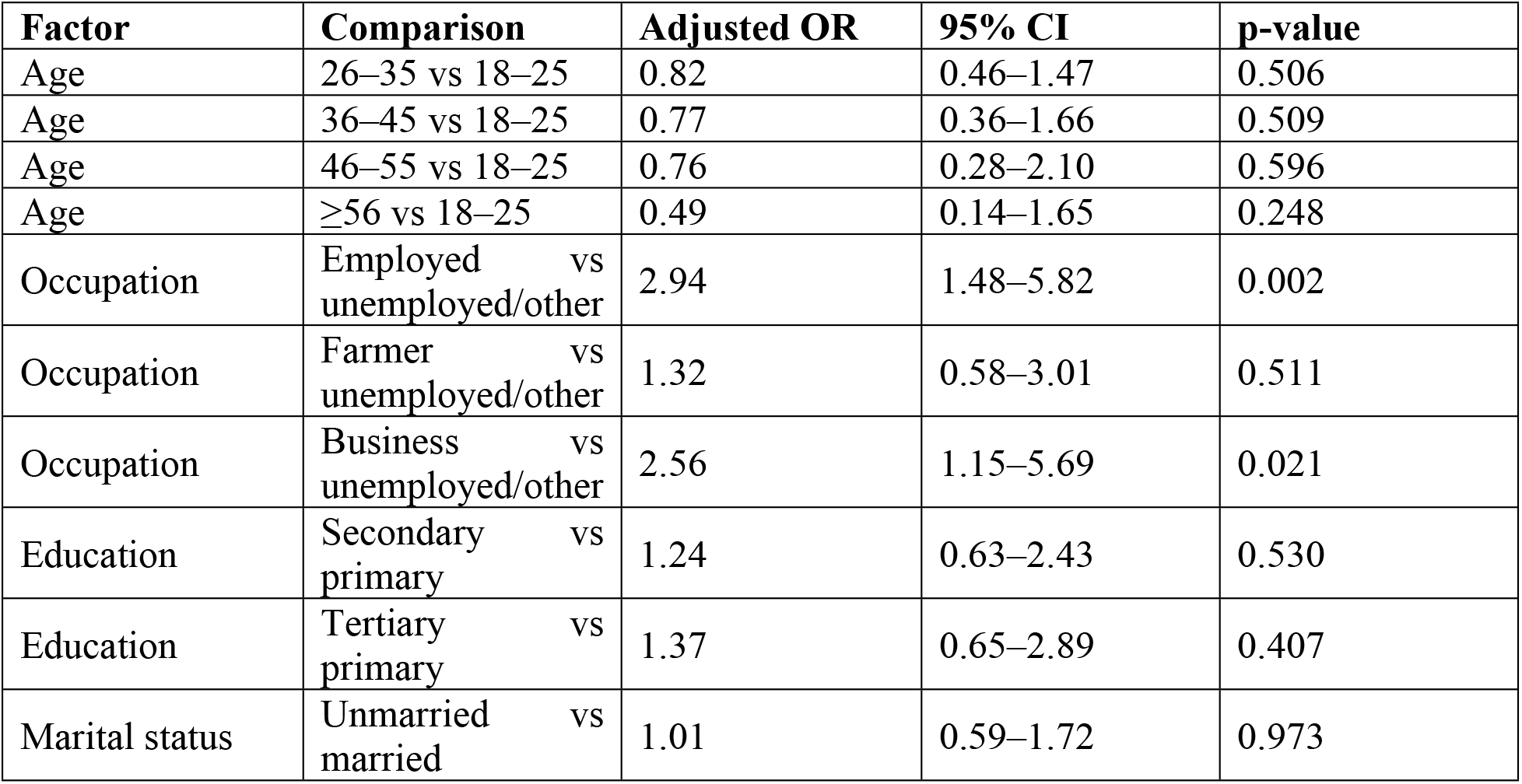
Adjusted socio-demographic factors associated with community-post HIV-testing utilization.

### Behavioral factors associated with community-post utilization

Among the 397 participants, 383 (96.5%) rated HIV testing as very important. Multiple sexual partnerships were reported by 235/397 (59.2%), unprotected sex by 221/397 (55.7%), alcohol or drug use by 183/397 (46.1%), peer influence by 83/397 (20.9%), injection-drug use by 39/397 (9.8%), and frequent travel or migration by 17/397 (4.3%). HIV testing was discussed frequently with friends, family or peers by 221 participants, occasionally by 104, and never by 70; two responses were missing.

In the adjusted behavioral model (complete-case n=391), respondents who were unwilling or unsure about participating in future community HIV-testing campaigns had lower odds of community-post use than those willing to participate (AOR 0.20, 95% CI 0.10–0.39; p<0.001). Frequent travel or migration was associated with lower odds of community-post use (AOR 0.19, 95% CI 0.06–0.65; p=0.008), as was reporting unprotected sex (AOR 0.54, 95% CI 0.34–0.87; p=0.012). Frequency of discussing HIV testing, perceived importance of testing, multiple sexual partnerships, injection-drug use, alcohol or drug use and peer influence were not independently associated with community-post utilization at the 5% level.

**Table 3.** Adjusted behavioral factors associated with community-post HIV-testing utilization.

| Factor | Comparison | Adjusted OR | 95% CI | p-value |
| --- | --- | --- | --- | --- |
| Discussion of HIV testing | Never vs frequently | 1.74 | 0.89–3.41 | 0.104 |
| Discussion of HIV testing | Occasionally vs frequently | 0.89 | 0.53–1.51 | 0.665 |
| Future campaign participation | No/Maybe vs yes | 0.20 | 0.10–0.39 | <0.001 |
| Multiple sexual partners | Yes vs no | 1.50 | 0.95–2.35 | 0.079 |
| Unprotected sex | Yes vs no | 0.54 | 0.34–0.87 | 0.012 |
| Injection-drug use | Yes vs no | 0.92 | 0.41–2.07 | 0.849 |
| Frequent travel/migration | Yes vs no | 0.19 | 0.06–0.65 | 0.008 |

| **Factor** | **Comparison** | **Adjusted OR** | **95% CI** | **p-value** |
| --- | --- | --- | --- | --- |
| Alcohol/drug use | Yes vs no | 1.00 | 0.62–1.61 | 0.996 |
| Peer influence | Yes vs no | 1.69 | 0.90–3.18 | 0.103 |

### Qualitative findings

Four recurrent themes emerged from the key informant interviews: accessibility and convenience, confidentiality and stigma, behavioral and social influences, and provider/service-delivery factors. Informants described community posts as easier for men to use because they were closer to where men worked or socialized, had shorter queues, and could operate beyond conventional facility hours. One informant stated, “The timing is the biggest difference. CPs are open until 7:00 pm, which is convenient for men after work” (KII 2). Confidentiality was also repeatedly emphasized. As another informant explained, “The key difference is the confidentiality. CPs are not solely for PLHIV, so the stigma of being seen there is lower” (KII 3). Fear of a positive HIV result and stigma were described as barriers to testing, alongside perceived risk, peer support, alcohol use, work demands and mobility. One informant summarized this concern: “Fear of positive results and associated stigma is the number one barrier. It outweighs awareness” (KII 1). Provider-related themes included staff attitudes, confidentiality, staffing levels, service quality and the availability of integrated care. Suggested improvements included increasing the number of community posts, regular staff training, maintaining confidentiality, and integrating services for other health needs so that community posts are not perceived as HIV-only service points. Numeric estimates offered by individual informants about changes in testing volumes were treated as perceptions and were not used as study prevalence or effect estimates because the underlying programme records were not available for verification.

## Discussion

This study examined factors associated specifically with men’s reported use of community posts for HIV testing in the preceding year. In the verified main dataset, just over half of respondents with complete outcome data reported community-post use (208/396; 52.5%). Because participants were recruited from HIV-testing service points and all reported at least one test in the preceding year, this proportion should be interpreted as the share reporting community-post use within the study sample, not as population-level HIV-testing coverage among men in Kericho County.

Occupation was independently associated with community-post utilization. Employed men and men engaged in business had higher adjusted odds of reporting community-post use than those classified as unemployed/other, while farming was not independently associated. The qualitative findings provide a plausible service-delivery context for this association. Informants emphasized opening hours, proximity to workplaces and social spaces, and shorter queues as features that could make community posts easier to use around work commitments. The statement that community posts could remain open until 7:00 pm was particularly relevant to men who worked during conventional facility hours. These accounts are consistent with, but do not establish, an explanation for the observed occupational differences. Age, marital status, religion, education and monthly income were not independently associated with community-post use in the adjusted model.

Selected behavioral factors were associated with community-post utilization after adjustment. Men who were unwilling or unsure about participating in future community HIV-testing campaigns had lower adjusted odds of reporting community-post use. Frequent travel or migration and unprotected sex were also associated with lower adjusted odds of community-post use. The interviews add context to these findings rather than confirming a causal pathway. Informants described fear of a positive result and stigma as important deterrents, while perceived HIV risk and supportive male peers were described as potential motivators. They also identified work and mobility as practical constraints to service use. These accounts show that behavioral decisions occurred alongside social and service-delivery considerations, but the cross-sectional data do not establish the direction, temporality or mechanism of the quantitative associations.

Other behavioral variables, including frequency of discussing HIV testing, multiple sexual partnerships, injection-drug use, alcohol or drug use and peer influence, were not independently associated with community-post utilization in the adjusted model.

The qualitative findings on confidentiality, clinic hours, stigma and fear of a positive result closely reflect evidence from Kenyan men. Okal and colleagues identified facility location, waiting time, inconvenient clinic hours, stigma, low perceived risk, fear of testing and provider attitudes as barriers, while confidentiality and flexible opening hours facilitated testing [7]. In the present study, informants similarly described extended hours and lower visibility of HIV-specific attendance as advantages of community posts. They also identified staff training, confidentiality, the number of available community posts and integration of other health services as areas for improvement. One informant argued that community posts should provide services such as hypertension or diabetes care so that the post is not viewed only as a place for HIV services. These findings situate the quantitative associations within the day-to-day service conditions described by providers in Kericho County.

### Strengths and limitations

The study combined quantitative and qualitative data from community and facility settings, allowing statistical associations to be interpreted alongside provider accounts of men’s service use. The cross-sectional design does not establish temporality or causality. Behavioral measures and testing locations were self-reported and may be affected by recall or social-desirability bias.

Participants could report more than one HIV-testing location, so service-location percentages are not mutually exclusive. One participant had a missing community-post outcome, and the behavioral multivariable model used complete cases. The qualitative component represents the experiences and perceptions of a small number of key informants and should not be used to estimate the frequency of behaviors or service outcomes.

## Conclusions

Among men in this study, community posts were a commonly reported HIV-testing location, with 208/396 (52.5%) of respondents with complete outcome data reporting community-post use in the preceding year. Employment and engagement in business were associated with higher adjusted odds of community-post use. Unwillingness or uncertainty about future community testing campaigns, frequent travel or migration, and unprotected sex were associated with lower adjusted odds. These associations identify groups and behaviors that may be considered when designing male-responsive community HIV-testing services, but causal conclusions cannot be drawn from the cross-sectional data.

## Data Availability

Data is available on KOBO tool extract and can be availed if needed

## Acknowledgments

The authors thank the study participants, research team, Kericho County Referral Hospital, Londiani Sub-County Hospital, Nyagacho Community Post and Brooke Community Post for their participation and support during the study.

## Author contributions

Lilian N. Kong’ani: Conceptualization, Methodology, Investigation, Data curation, Formal analysis, Writing – original draft, Writing – review & editing. Jackline Mosinya Nyaberi: Supervision, Methodology, Formal analysis, Writing – review & editing. Kenneth Ngure: Supervision, Conceptualization, Methodology, Interpretation of results, Writing – review & editing.

